# Antibody Responses to SARS-CoV-2 in Coronavirus Diseases 2019 Patients with Different Severity

**DOI:** 10.1101/2020.09.06.20189480

**Authors:** Ekasit Kowitdamrong, Thanyawee Puthanakit, Watsamon Jantarabenjakul, Eakachai Prompetchara, Pintip Suchartlikitwong, Opass Putcharoen, Nattiya Hirankarn

## Abstract

**Background:** More understanding of antibody responses in the SARS-CoV-2 infected population is useful for vaccine development.

**Aim:** To investigate SARS-CoV-2 IgA and IgG among COVID-19 Thai patients with different severity.

**Methods:** We used plasma from 118 adult patients who have confirmed SARS-CoV-2 infection and 49 patients under investigation without infection, 20 patients with other respiratory infections, and 102 healthy controls. Anti-SARS-CoV-2 IgA and IgG were performed by enzyme-linked immunosorbent assay from Euroimmun. The optical density ratio cut off for positive test was 1.1 for IgA and 0.8 for IgG. The association of antibody response with the severity of diseases and the day of symptoms was performed.

**Results:** From Mar 10 to May 31, 2020, 289 participants were enrolled, and 384 samples were analyzed. Patients were categorized by clinical manifestations to mild (n = 59), moderate (n = 27) and severe (n = 32). The overall sensitivity of IgA and IgG from samples collected after day 7 is 87.9% (95% CI 79.8-93.6) and 84.8% (95% CI 76.2-91.3), respectively. The severe group had a significantly higher level of specific IgA and IgG to S1 antigen compared to the mild group. All moderate to severe patients have specific IgG while 20% of the mild group did not have any IgG detected after two weeks. Interestingly, SARS-CoV-2 IgG level was significantly higher in males compared to females among the severe group (p = 0.003).

**Conclusion:** The serologic test for SARS-CoV-2 has high sensitivity after the second week after onset of illness. Serological response differs among patients with different severity and different sex.

## Introduction

In late December 2019, an outbreak with initially undiagnosed pneumonia was reported in Wuhan, Hubei Province, China. [1]. The causative pathogen was later identified as a novel beta coronavirus closely related to severe acute respiratory syndrome (SARS) coronavirus (CoV) family, recently termed SARS-CoV-2 [2]. As of 30^th^ July 2020, more than 17 million people infected with SARS-CoV-2 and up to 670,000 death tolls [3]. The first case in Thailand was reported on Jan 12, a traveler from Wuhan to Thailand [4]. As of 30^th^ July 2020, there were 3,304 cases confirmed in Thailand with the epicenter in Bangkok metropolitan area. Real-time RT-PCR diagnostic assays is a goal standard for case ascertainment and diagnosis [5]. However, validated serologic tests are evidence to compliment virological diagnosis, particularly in the second week of infection [6]. More understanding of antibody responses in the infected population will be useful for vaccine development.

The ELISA is commonly used to access viral-specific antibodies in quantitative manners and is widely accepted as an antibody diagnostic test for decades. The sensitive, quantitative measure of ELISA makes it suitable to assess the dynamic change of the viral-specific antibodies. In principle, specific IgM and IgA should be detected first approximately in the second week of infection followed by specific IgG after the second week of infection. There are several serology platforms available now using various antigens. One large nucleocapsid-based ELISA study of 208 samples reported that the IgM and IgA were detected at 3-6 days after symptoms with the sensitivity of 85.4 and 92.7 %, respectively; while IgG was detected later at 10-18 days post-infection with the sensitivity of 77.9 % [7]. Interestingly, another study showed that IgG seroconversion against SARS-CoV-2 nucleocapsid and a peptide from spike region was detected as early as that of IgM and reached its peak within six days after seroconversion [8]. A weaker and more rapid decline of antibody response was observed in asymptomatic and patients with milder symptoms compared to severe patients [9].

The Euroimmun Anti-SARS-CoV-2 enzyme-linked immunosorbent assay (ELISA) was one of the first CE-marked diagnostic assay developed and available worldwide. It assesses the IgA and IgG to the spike 1 (S1) protein and has been reported to correlate well with plaque reduction neutralization test (PRNT) [10–11]. The Euroimmun IgG assay has received EUA from the US Food and Drug Administration. So far, most of the results were reported from Europe and the USA. The objective of this study is to investigate the dynamics of IgA and IgG antibody against SARS-CoV-2 in serial blood samples collected from confirmed COVID-19 patients in the Thai population and associated with severity of illness.

## Materials and Methods

The study was conducted at the Thai Red Cross Emerging Infectious Diseases Clinical Center (TRC-EIDCC) and Faculty of Medicine, Chulalongkorn University. The study was reviewed and approved by the Institutional Review Board of Faculty of Medicine (IRB number 242/63) and National Blood Center, Thai Red Cross Society (COA No. NBC 5/2020).

## Patient population

Confirmed COVID-19 cases were defined as testing positive for SARS-CoV-2 RNA using real-time reverse transcription-polymerase chain reaction (RT-PCR) testing from combined nasopharyngeal and throat swab (NT) samples at Department of Microbiology, Faculty of Medicine, Chulalongkorn University. SARS-CoV-2 RNA was detected using cobas® SARSCoV-2 kit (Roche Diagnostics, Switzerland) on the fully automated cobas® 6800 system (Roche Diagnostics, Switzerland) according to the manufacturer’s recommendation. Nucleic acid was automatically extracted from 400 microliters of NT in viral transport medium (VTM) along with added internal control RNA (RNA IC). Subsequent real-time RT-PCR was done automatically by the system targeting ORF1a/b and E genes specific to SARS-CoV-2 and pan-Sarbecovirus, respectively.

Classification of the confirmed case was 1) mild – asymptomatic or upper respiratory tract infection 2) moderate – pneumonia without hypoxia 3) severe – pneumonia with hypoxia. The date of illness onset, disease severity, the hospitalization period, the personal demographic information was obtained from the hospital medical records. The Control group included three groups. The first group was 20 plasma from healthy volunteers in the laboratory and 82 leftover plasma samples collected from blood donor healthy volunteers before February 2020. The second group was 49 plasma samples from the patient under investigation (PUI) for COVID-19 but had RT-PCR negative for SARS-CoV-2 collected during May 1 to May 31, 2020. A third control group was 20 serum specimens collected from patients with other infections (Dengue, HBV, HCV, HIV, Mumps, Measles, Rubella, EBV, CMV, VZV, HSV, Treponema) collected during May 1 to May 31, 2020. Plasma and serum were aliquot and kept at −20 °C before carrying out serologic tests.

## Laboratory methods

Ten microliters of plasma samples were diluted to 1:101 in sample buffer in order to perform SARS-CoV-2 S1-specific IgA and IgG ELISAs using Anti-SARS-CoV-2 ELISA (IgG) and Anti-SARS-CoV-2 ELISA (IgA) kits (Euroimmun, Germany) according to the manufacturer’s instructions. Semiquantitative results were evaluated by calculating a ratio of the extinction at 450 nm of each sample over the calibrator. The cutoff ratio of 1.1 was used for SARS-CoV-2 IgA, as suggested by the package insert. The borderline cutoff ratio of 0.8 for SARS-CoV-2 IgG was assigned as positive.

## Statistical analysis

Demographic was described for the patient. Continuous variables are expressed as median (interquartile range: IQR). Differences in continuous and categorical variables between the two groups were assessed using a Wilcoxon rank-sum test and Chi-square test or Fisher exact test, respectively. Sensitivity, specificity, positive predictive value (PPV) and negative predictive value (NPV) were calculated.

## Results

### Demographics of the population

From Mar 10 to May 31, 2020, there were 118 confirmed SARS-CoV-2 infection; 59 with mild (upper respiratory symptoms), 27 with moderate (pneumonia), 32 with severe (pneumonia with hypoxia), with a median age of 38 years (IQR 27-48). A total of 213 samples collected from the 118 patients were tested for antibodies against SARS-Co-V2. The number of second samples (N = 82), 3 samples (n = 13). A total of 99 samples were collected after at least seven days from the onset of illness. Forty-nine patients under investigation had negative SARS-CoV-2 with a median age of 47 years (IQR 28-65 years), with 25 males and 24 females. Baseline clinical characteristics were summarized in table 1. In summary, there was significantly different in age and sex between groups. The severe patients were mostly male (66%) and within the 40-59 age group.

**Table 1:** Clinical characteristics of patients

|  | COVID<br>(N=118) | Mild<br>(N=59) | Mod<br>(N=27) | Severe<br>(N=32) | Non-COVID<br>(N=49) | P-value |
| --- | --- | --- | --- | --- | --- | --- |
| Median Age<br>(IQR) | 38 (27-48) | 29 (26-39) | 39 (27-47) | 49 (41-58) | 47 (28-65) | <0.001 |
| Age group, n (%) |  |  |  |  |  | <0.001 |
| • < 20 | 6 (5) | 4 (7) | 1 (4) | 1 (3) | 3 (6) |  |
| • 20-39 | 61 (52) | 41 (70) | 14 (52) | 6 (19) | 17 (35) |  |
| • 40-59 | 43 (36) | 12 (20) | 11 (40) | 20 (62) | 14 (28) |  |
| • > 60 | 8 (7) | 2 (3) | 1 (4) | 5 (16) | 15 (31) |  |
| Male, n (%) | 47 (40) | 19 (32) | 7 (26) | 21 (66) | 25 (51) | <0.001 |

### Seroconversion of antibodies against SARS-CoV-2 in COVID 19 Patients

Among 118 confirmed SARS-CoV-2 infection, 99 patients had blood samples collected at least once after day 7 from onset of symptoms. The overall seroconversion of antibodies after day seven from the onset of illness was summarized in table 2. The overall sensitivity of IgA is 87.9% (95% CI 79.8-93.6) and negative predictive value 93.1% (95%CI 88.3-96.4). The overall sensitivity of IgG is 84.8% (95% CI 76.2-91.3) and negative predictive value 91.0% (95%CI 87.9-96.1). The overall specificity of IgA and IgG is 94.7% and 97.1%, respectively.

**Table 2.** The overall sensitivity of samples collected at day of symptoms > 7 days ELISA_IgA ≥ 1.1

| **ELISA IgA $\geq 1.1$** | **n/N** | **%** | **95%CI** |  |
| --- | --- | --- | --- | --- |
| Sensitivity | 87/99 | 87.9 | 79.8 | 93.6 |
| Specificity<sup>a</sup> | 162/171 | 94.7 | 90.2 | 97.6 |
| Positive predictive value | 87/96 | 90.6 | 82.9 | 95.6 |
| Negative predictive value | 162/174 | 93.1 | 88.3 | 96.4 |
| ROC area (Sens. + Spec.)/2 | - | 0.91 | 0.88 | 0.95 |
| **ELISA IgG $\geq 0.8$** |  | **%** | **95%CI** |  |
| Sensitivity | 84/99 | 84.8 | 76.2 | 91.3 |
| Specificity<sup>b</sup> | 166/171 | 97.1 | 93.3 | 99 |
| Positive predictive value | 84/89 | 94.4 | 87.4 | 98.2 |
| Negative predictive value | 166/181 | 91.7 | 87.9 | 96.1 |
| ROC area (Sens. + Spec.)/2 | - | 0.91 | 0.87 | 0.95 |

<sup>a</sup> IgA Specificity in Healthy control = 100/102 = 98.03%, Specificity in Patients under investigation for COVID-19 with SARS-CoV-2 RT-PCR negative = 44/49 = 89.8%, Specificity in cross-reactivity panel group = 18/20 = 90%
<sup>b</sup> IgG Specificity in Healthy control = 101/102 = 99.01%, Specificity in Patients under investigation for COVID-19 with SARS-CoV-2 RT-PCR negative = 47/49 = 95.9%, Specificity in cross-reactivity panel group = 18/20 = 90%

However, the specificity varies in a different group of control. The raw data of all control were shown in S1 table. There were 2 false-positive IgA and only one false positive IgG in 102 healthy control. We were able to obtain and re-analyzed seven samples with OD ratio ≥ 0.8 after two months interval. The OD ratio was quite similar to the initial result confirming the healthy control group’s little background. The higher background was observed in the control groups with respiratory symptoms. There were five positive IgA and two positive IgG in 49 patients under investigation for COVID-19 with negative RT-PCR for SARS-CoV-2. Two of these patients were repeated after 2-4 weeks, and the OD ratio was back to normal. It should be noted that the positive results in these patients might be the result of either false positive or true positive cases with negative RT-PCR. However, we did not have any evidence to support the COVID-19 infection in these patients. From 20 serum specimens collected from patients with other infections revealed two samples with both IgA and IgG cross-reactivity with CMV and EBV positive samples.

### Seroconversion of antibody stratifies by day of illness and disease severity

The seroconversion of antibodies stratifies by day of illness was shown in table 3. Sensitivity for a serologic test within seven days from onset of illness is only 29.7 – 30.6% for IgA and 10.2-16.2% for IgG. The IgA positive rate was increased to 60% during the 2^nd^ week and 100% during the 3-4th week, and then decline to 81.9% in the second month. The positive IgG rate was increased to 90% during the 3^rd^-4th week of diseases.

To investigate the antibody level according to the severity of the disease, the antibody levels at the first time point were expressed using the cutoff value stratify by disease severity. The severe group had a significantly higher level of specific IgA and IgG to S1 antigen compared to the mild group (Fig 1). It should be noted that the two patients in the severe group who did not have specific IgA detected were tested only one time at 31- and 40-days post symptom. It is likely that IgA was already declined in these patients.

**Figure 1.**
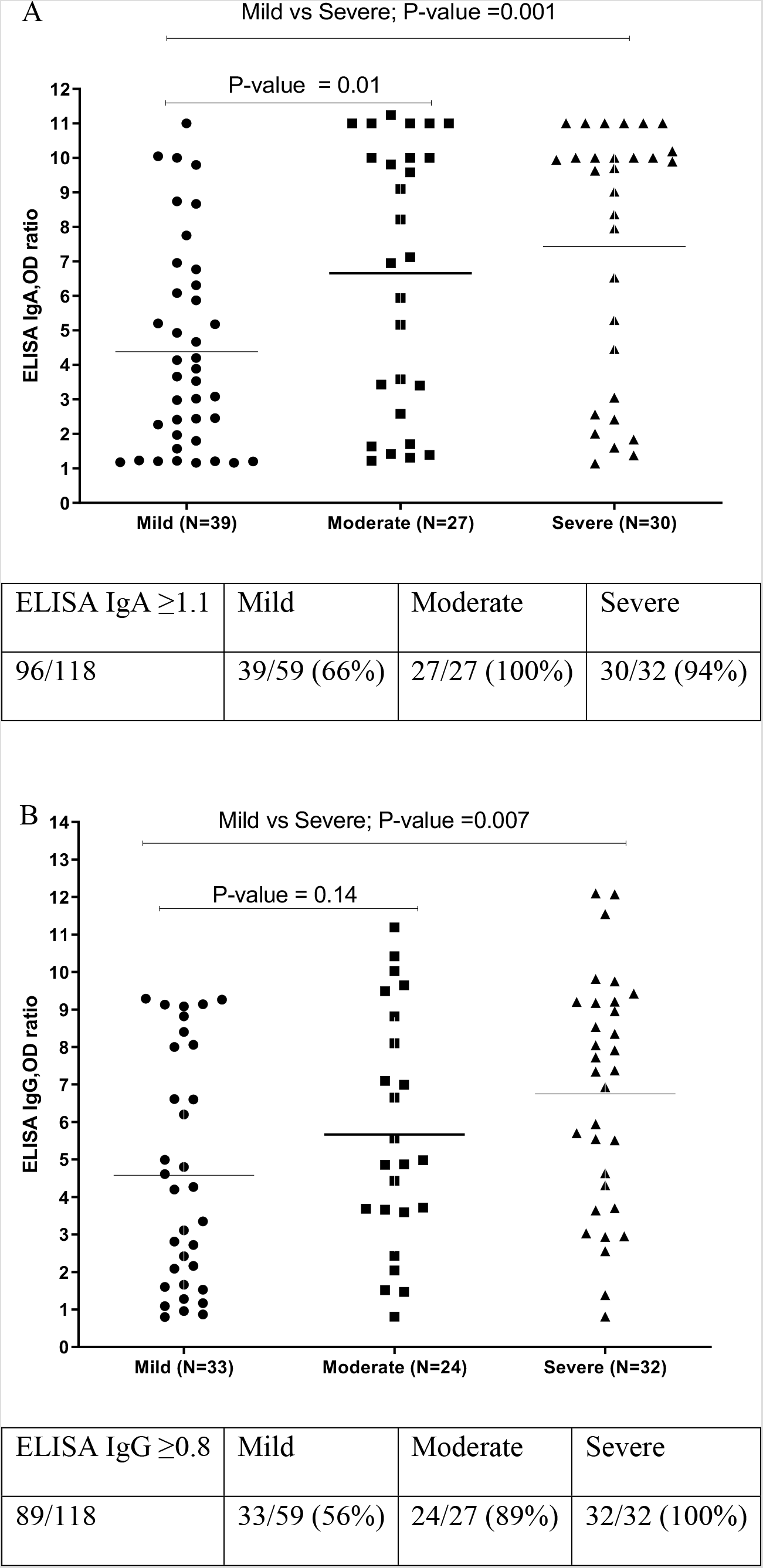
Antibody level according to the disease severity A) ELISA IgA OD ratio, B) ELISA IgG OD ratio

To see the dynamic from each group, we plot the average antibody level from mild, moderate, and severe groups at five intervals (Fig 2).

**Figure 2.**
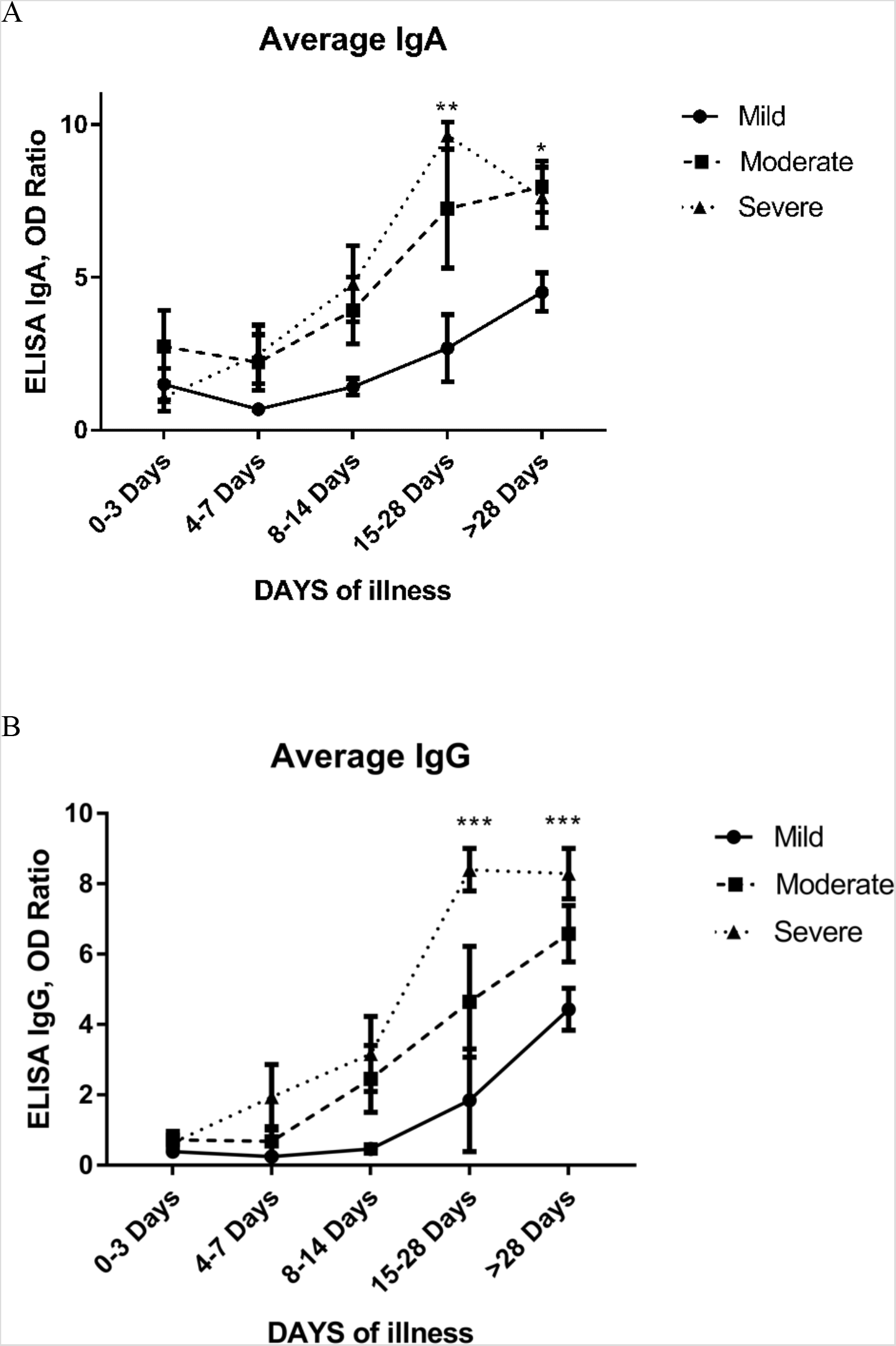
The average levels of A) IgA OD ratio and B) IgG OD ratio to SARS-CoV-2 among COVID-19 patients with different severity by days of illness

There were 103, 52, and 58 samples for mild, moderate, and severe groups, respectively (Table 3). A clear pattern shows that a severe and moderate group has significantly higher IgA and IgG after 15 days post symptoms compared to the mild group. There were 20% (7/35) of samples with mild symptoms that did not have IgG after the second week. Only 1 out of 15 patients with moderate symptoms did not have IgG while all 15 patients with severe symptoms had high IgG titer after the second week (Table 3).

**Table 3.** The seroconversion of antibody stratifies by day of illness and severity (N = 213 tests)

| | PCR positive | ELISA IgA $\geq 1.1$ | | | |
| --- | --- | --- | --- | --- | --- |
|  |  | Total | Mild | Moderate | Severe |
| Day 0-3 | 37 | 11/37 (29.7%) | 6/23 (26%) | 3/9 (33.3%) | 2/5 (40%) |
| Day 4-7 | 49 | 15/49 (30.6%) | 3/25 (12%) | 7/10 (70%) | 5/14 (35.7%) |
| Day 8-14 | 45 | 27/45 (60%) | 8/20 (40%) | 9/13 (69.2%) | 10/12 (83.3%) |
| Day 15-28 | 21 | 21/21 (100%) | 4/4 (100%) | 5/5 (100%) | 12/12 (100%) |
| Day > 28 | 61 | 50/61 (81.9%) | 22/31 (71%) | 14/15 (93.3%) | 14/15 (93.3%) |
|  | 213 | 124/213 (58.2%) | 43/103 (41.7%) | 38/52 (73.1%) | 43/58 (74.1%) |
| | PCR positive | ELISA IgG $\geq 0.8$ | | | |
|  |  | Total | Mild | Moderate | Severe |
| Day 0-3 | 37 | 6 (16.2%) | 1/23 (4.3%) | 3/9 (33.3%) | 2/5 (40%) |
| Day 4-7 | 49 | 5 (10.2%) | 0/25 (0%) | 1/10 (10%) | 4/14 (28.6%) |
| Day 8-14 | 45 | 14 (31.1%) | 4/20 (20%) | 4/13 (30.8%) | 6/12 (50%) |
| Day 15-28 | 21 | 19 (90.5%) | 2/4 (50%) | 5/5 (100%) | 12/12 (100%) |
| Day > 28 | 61 | 55 (90.2%) | 26/31 (83.9%) | 14/15 (93.3%) | 15/15 (100%) |
|  | 213 | 99 (46.5%) | 33/103 (32%) | 27/52 (51.9%) | 39/58 (67.2%) |

Since age and sex were associated with disease outcome, we analyzed the correlation between antibody titer and age in the severe group, as shown in Fig3A and B. There was no significant correlation.

**Figure 3.**
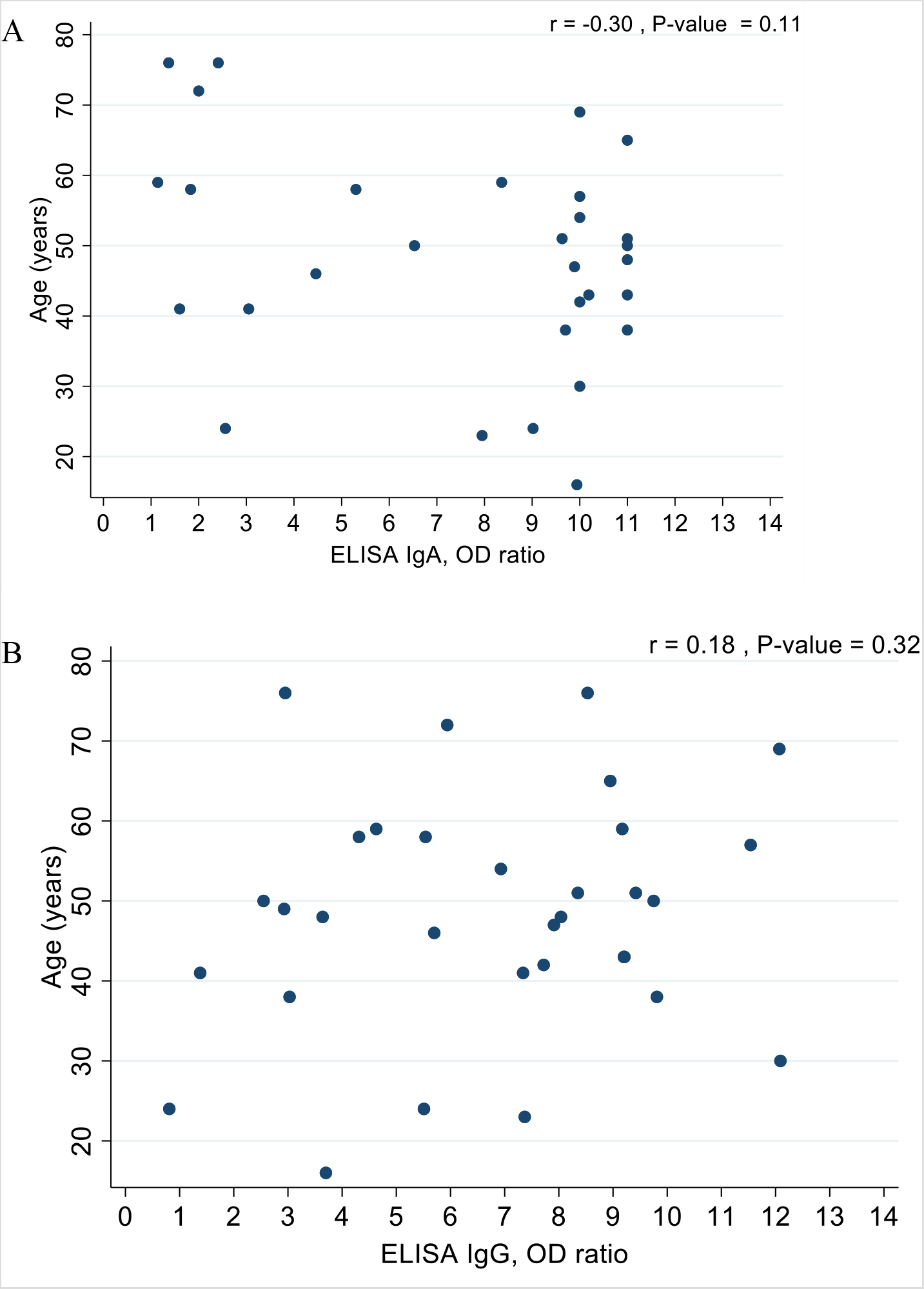
The correlation between Ab titer and Age in severe group A) Age VS ELISA IgA OD ratio, B) Age VS ELISA IgG OD ratio

We also compared antibody titer between males and females within the severe group. Interestingly, the level of both IgA and IgG to S1 antigen was higher in males than in females. Mainly IgG was statistically significant (Fig 4A and 4B). The median age of male (51 with IQR = 43-59) was also higher than female (41 with IQR = 24-46) among the severe group.

**Figure 4.**
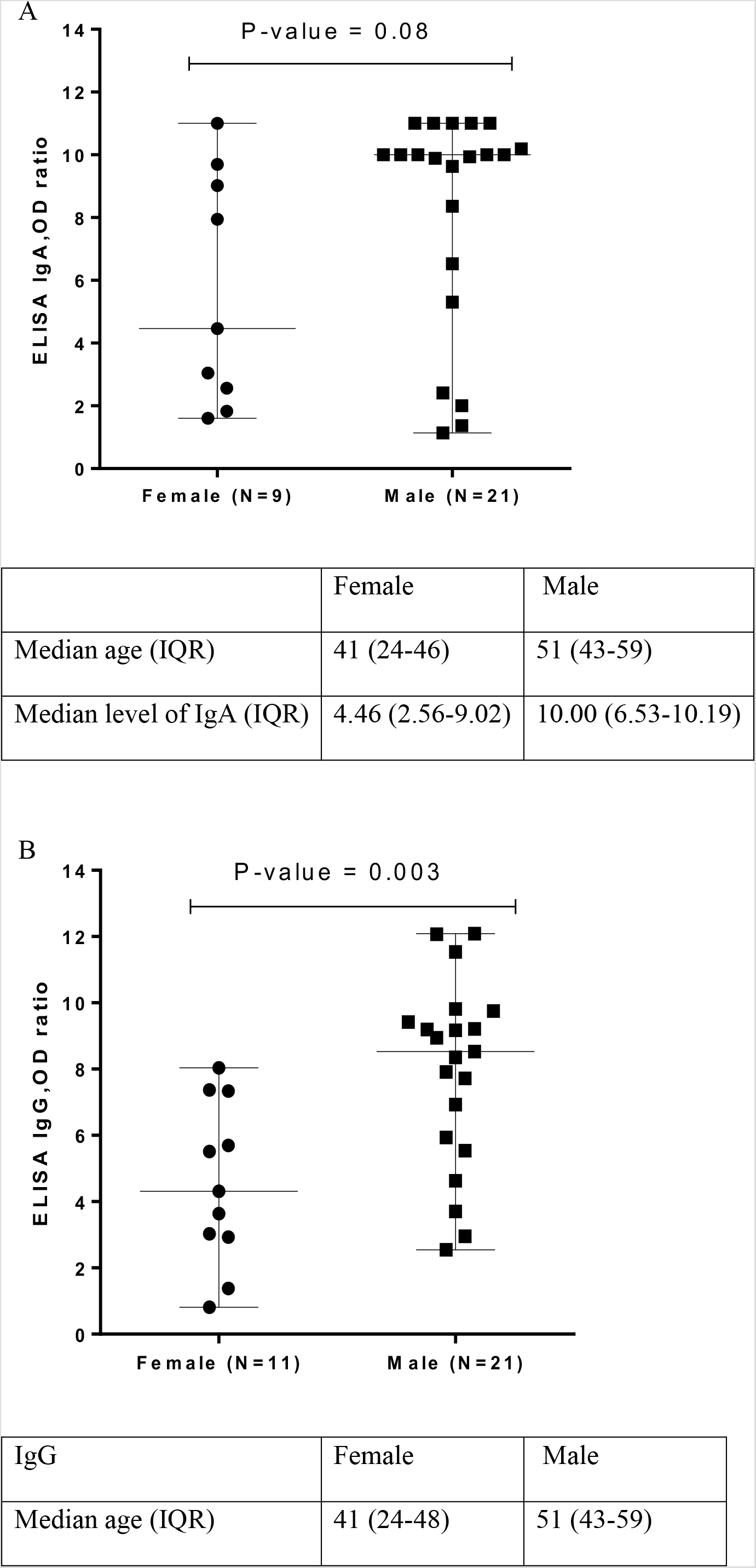
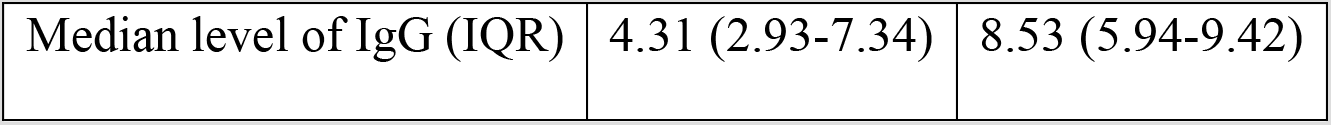
The relationship between Ab titer and Sex in severe group A) Sex VS ELISA IgA OD ratio, B) Sex VS ELISA IgG OD ratio

## Discussion

The data demonstrated an antibody response to acute viral infection that the sensitivity in the first week of infection is low. The antibody response shown in this study was IgA and then followed by IgG. Since it is difficult to compare results using different serological analysis, we will only use data from studies that tested with Euroimmun for comparison. The results from 15 studies based on Euroimmun were summarized in S2 table. Previous studies mostly reported that the sensitivity of IgA within the first week was less than 60% [12–13]. There was 30% of COVID-19 patients who developed positive IgA very early within 3 days after the symptom in this study. Therefore, positive IgA might help identify some COVID-19 patients in the early phase, but the negative result cannot be used to exclude the infection. The seroconversion of IgA was 100% in the 21 patients at 15-28 days after the symptoms. A hundred percent sensitivity of IgA seroconversion was also reported in 82 cases from France after the second weeks of disease onset [13] and 91 patients after the third weeks [14]. Interestingly, we noticed the decline of IgA after one month. The sensitivity of IgA after one month was decreased to 80% in this study.

As for the IgG, it should be noted that we used the borderline cut off as a positive result in this study to increase the sensitivity of IgG. This cut off did not change the specificity of the test. The IgG specific to SARS-CoV-2 S1 antigen occurs later than IgA. The sensitivity of IgG was 90% after the second week of disease onset. This number is quite comparable to other reports [13, 15–17].

In this study, 20% of the patients with mild symptoms did not mount any specific IgG to the virus even after 2 weeks. Other reports found up to 20-30% negative IgG [18–19]. When we analyzed the correlation of antibody titer with clinical severity, it was clear that patients with more severe clinical manifestation had a higher antibody titer both, IgA and IgG than, patients in the mild group. This observation is consistently reported in other populations as well [10, 19, 20]. The explanation of these findings is not clear yet. There is a hypothesis that the higher inflammatory environment in the severe patients might induce a more robust immune response, including the antibody production from the B lymphocytes. It also raises concerns about the role of antibody-mediated severity, although there is no evidence to support it. Besides, several studies reported that there was a higher rate of severity and mortality in male patients [21]. In our cohort, more females are infected with COVID-19 than male patients in total (60% female VS 40% male). However, there was significantly more male (66%) in the severe group. Interestingly, we found a significantly higher level of IgG in males than females among the severe groups, similar to recent studies from Klein et al. [19]. The median age of the males was higher than females among the severe group too. It is possible that higher level of antibody might simply associate with more severity in the male patients. However, there is a speculation that biological sex might affect immunity by various mechanisms [21]. Although women seem to have greater antibody responses and are susceptible to autoimmune diseases than men, other factors, including innate immunity, regulatory T cells, expression of angiotensin-converting enzyme 2, or other factors relate to sex hormone might explain the more severity and high antibody titer observed in male patients. More studies to unravel the role of sex impact on disease severity might lead to a better understanding of this challenging disease.

In summary, this study extensively reported the serological responses of COVID-19 patients in Thailand up to 60 days after the disease onset. Although most of the samples were tested at two time points, the blood samples were collected from patients at different stages and at various intervals. Therefore, we did not perform a median time of positive, which might be subjected to bias.

## Data Availability

All the data was presented in the manuscript.

## Acknowledgements

This work was supported by funding to support Biobank from Ratchadapisek Sompoch Fund, Faculty of Medicine, Chulalongkorn University. We would like to thank the health care team for at King Chulalongkorn Memorial hospital, Thai Red Cross, particularly Dr. Kampol Suwanpimolkul, Dr. Leilanee Paitoonpong, and Dr. Suvaporn Anulgulreungkitt. Special thanks for the advice from Dr. Parvapan Bhattarakosol and statistical analysis by Miss Jiratchaya Sophonphan.

## Supporting information

**S1 Table:** Raw data of control group

| No | Details | ELISA_Ig<br>A<br>(OD<br>Ratio) | Interpretati<br>on (0=neg,<br>1=pos) | ELISA_Ig<br>G (OD<br>Ratio) | Interpretati<br>on (0=neg,<br>1=pos) |  | ELISA_Ig<br>A (OD<br>Ratio) | ELISA_Ig<br>G (OD<br>Ratio) |
| --- | --- | --- | --- | --- | --- | --- | --- | --- |
| 1 | Healthy<br>donor | 0.57 | 0 | 0.13 | 0 |  |  |  |
| 2 | Healthy<br>donor | 0.47 | 0 | 0.18 | 0 |  |  |  |
| 3 | Healthy<br>donor | 0.39 | 0 | 0.19 | 0 |  |  |  |
| 4 | Healthy<br>donor | 0.69 | 0 | 0.19 | 0 |  |  |  |
| 5 | Healthy<br>donor | 0.12 | 0 | 0.21 | 0 |  |  |  |
| 6 | Healthy<br>donor | 1.32 | 1 | 0.18 | 0 | Repea<br>t after<br>2<br>month<br>s | 1.26 | 0.19 |
| 7 | Healthy<br>donor | 0.44 | 0 | 0.52 | 0 |  |  |  |
| 8 | Healthy<br>donor | 0.26 | 0 | 0.17 | 0 |  |  |  |
| 9 | Healthy<br>donor | 0.88 | 0 | 0.35 | 0 | Repea<br>t after<br>2<br>month<br>s | 0.97 | 0.37 |
| 10 | Healthy<br>donor | 0.29 | 0 | 0.17 | 0 |  |  |  |
| 11 | Healthy<br>donor | 0.39 | 0 | 0.17 | 0 |  |  |  |
| 12 | Healthy<br>donor | 0.40 | 0 | 0.17 | 0 |  |  |  |
| 13 | Healthy<br>donor | 0.33 | 0 | 0.14 | 0 |  |  |  |
| 14 | Healthy<br>donor | 0.80 | 0 | 0.23 | 0 | Repea<br>t after<br>2<br>month<br>s | 0.75 | 0.22 |
| 15 | Healthy<br>donor | 0.95 | 0 | 0.16 | 0 | Repea<br>t after<br>2<br>month<br>s | 0.97 | 0.17 |
| 16 | Healthy<br>donor | 0.80 | 0 | 0.13 | 0 | Repea<br>t after<br>2<br>month<br>s | 0.84 | 0.19 |
| 17 | Healthy<br>donor | 0.22 | 0 | 0.12 | 0 |  |  |  |
| 18 | Healthy<br>donor | 0.23 | 0 | 0.22 | 0 |  |  |  |
| 19 | Healthy donor | 0.83 | 0 | 0.22 | 0 | Repeat after 2 months | 0.92 | 0.31 |
| 20 | Healthy donor | 0.66 | 0 | 0.23 | 0 |  |  |  |
| 21 | Healthy donor | 0.58 | 0 | 0.19 | 0 |  |  |  |
| 22 | Healthy donor | 0.45 | 0 | 0.21 | 0 |  |  |  |
| 23 | Healthy donor | 0.69 | 0 | 0.21 | 0 |  |  |  |
| 24 | Healthy donor | 0.45 | 0 | 0.15 | 0 |  |  |  |
| 25 | Healthy donor | 0.20 | 0 | 0.21 | 0 |  |  |  |
| 26 | Healthy donor | 0.28 | 0 | 0.15 | 0 |  |  |  |
| 27 | Healthy donor | 0.64 | 0 | 0.23 | 0 |  |  |  |
| 28 | Healthy donor | 0.34 | 0 | 0.14 | 0 |  |  |  |
| 29 | Healthy donor | 0.35 | 0 | 0.14 | 0 |  |  |  |
| 30 | Healthy donor | 0.53 | 0 | 0.52 | 0 |  |  |  |
| 31 | Healthy donor | 0.40 | 0 | 0.17 | 0 |  |  |  |
| 32 | Healthy donor | 0.92 | 0 | 0.36 | 0 | Repeat after 2 months | 1.11 | 0.44 |
| 33 | Healthy donor | 0.47 | 0 | 0.15 | 0 |  |  |  |
| 34 | Healthy donor | 0.28 | 0 | 0.21 | 0 |  |  |  |
| 35 | Healthy donor | 0.28 | 0 | 0.15 | 0 |  |  |  |
| 36 | Healthy donor | 0.27 | 0 | 0.17 | 0 |  |  |  |
| 37 | Healthy donor | 0.36 | 0 | 0.15 | 0 |  |  |  |
| 38 | Healthy donor | 0.78 | 0 | 0.15 | 0 |  |  |  |
| 39 | Healthy donor | 0.37 | 0 | 0.15 | 0 |  |  |  |
| 40 | Healthy donor | 0.18 | 0 | 0.21 | 0 |  |  |  |
| 41 | Healthy donor | 0.24 | 0 | 0.15 | 0 |  |  |  |
| 42 | Healthy donor | 0.47 | 0 | 0.31 | 0 |  |  |  |
| 43 | Healthy donor | 0.46 | 0 | 0.16 | 0 |  |  |  |
| 44 | Healthy donor | 0.32 | 0 | 0.18 | 0 |  |  |  |
| 45 | Healthy donor | 0.27 | 0 | 0.16 | 0 |  |  |  |
| 46 | Healthy donor | 0.25 | 0 | 0.14 | 0 |  |  |  |
| 47 | Healthy donor | 0.37 | 0 | 0.24 | 0 |  |  |  |
| 48 | Healthy donor | 0.87 | 0 | 0.26 | 0 |  |  |  |
| 49 | Healthy donor | 0.42 | 0 | 0.19 | 0 |  |  |  |
| 50 | Healthy donor | 0.06 | 0 | 0.24 | 0 |  |  |  |
| 51 | Healthy donor | 0.25 | 0 | 0.22 | 0 |  |  |  |
| 52 | Healthy donor | 0.78 | 0 | 0.42 | 0 |  |  |  |
| 53 | Healthy donor | 0.26 | 0 | 0.21 | 0 |  |  |  |
| 54 | Healthy donor | 0.30 | 0 | 0.14 | 0 |  |  |  |
| 55 | Healthy donor | 0.42 | 0 | 0.18 | 0 |  |  |  |
| 56 | Healthy donor | 0.25 | 0 | 0.18 | 0 |  |  |  |
| 57 | Healthy donor | 0.37 | 0 | 0.17 | 0 |  |  |  |
| 58 | Healthy donor | 0.42 | 0 | 0.18 | 0 |  |  |  |
| 59 | Healthy donor | 0.61 | 0 | 0.44 | 0 |  |  |  |
| 60 | Healthy donor | 0.46 | 0 | 0.17 | 0 |  |  |  |
| 61 | Healthy donor | 0.51 | 0 | 0.20 | 0 |  |  |  |
| 62 | Healthy donor | 0.45 | 0 | 0.13 | 0 |  |  |  |
| 63 | Healthy donor | 0.34 | 0 | 0.14 | 0 |  |  |  |
| 64 | Healthy donor | 0.20 | 0 | 0.18 | 0 |  |  |  |
| 65 | Healthy donor | 0.78 | 0 | 0.17 | 0 |  |  |  |
| 66 | Healthy donor | 0.42 | 0 | 0.24 | 0 |  |  |  |
| 67 | Healthy donor | 0.37 | 0 | 0.16 | 0 |  |  |  |
| 68 | Healthy donor | 0.28 | 0 | 0.13 | 0 |  |  |  |
| 69 | Healthy donor | 2.83 | 1 | 0.50 | 0 |  |  |  |
| 70 | Healthy donor | 0.24 | 0 | 0.35 | 0 |  |  |  |
| 71 | Healthy donor | 0.49 | 0 | 0.17 | 0 |  |  |  |
| 72 | Healthy donor | 0.22 | 0 | 0.17 | 0 |  |  |  |
| 73 | Healthy donor | 0.44 | 0 | 0.47 | 0 |  |  |  |
| 74 | Healthy donor | 0.29 | 0 | 0.27 | 0 |  |  |  |
| 75 | Healthy donor | 0.28 | 0 | 0.16 | 0 |  |  |  |
| 76 | Healthy donor | 0.56 | 0 | 0.20 | 0 |  |  |  |
| 77 | Healthy donor | 0.22 | 0 | 0.30 | 0 |  |  |  |
| 78 | Healthy donor | 0.12 | 0 | 0.12 | 0 |  |  |  |
| 79 | Healthy donor | 0.37 | 0 | 0.26 | 0 |  |  |  |
| 80 | Healthy donor | 0.68 | 0 | 0.13 | 0 |  |  |  |
| 81 | Healthy donor | 0.19 | 0 | 0.14 | 0 |  |  |  |
| 82 | Healthy donor | 0.51 | 0 | 0.20 | 0 |  |  |  |
| 83 | Healthy donor | 0.19 | 0 | 0.22 | 0 |  |  |  |
| 84 | Healthy donor | 0.26 | 0 | 0.17 | 0 |  |  |  |
| 85 | Healthy donor | 0.13 | 0 | 0.21 | 0 |  |  |  |
| 86 | Healthy donor | 0.25 | 0 | 0.16 | 0 |  |  |  |
| 87 | Healthy donor | 0.12 | 0 | 0.13 | 0 |  |  |  |
| 88 | Healthy donor | 0.20 | 0 | 0.16 | 0 |  |  |  |
| 89 | Healthy donor | 0.51 | 0 | 0.20 | 0 |  |  |  |
| 90 | Healthy donor | 0.30 | 0 | 0.32 | 0 |  |  |  |
| 91 | Healthy donor | 0.41 | 0 | 0.16 | 0 |  |  |  |
| 92 | Healthy donor | 0.26 | 0 | 0.28 | 0 |  |  |  |
| 93 | Healthy donor | 0.48 | 0 | 1.29 | 1 |  |  |  |
| 94 | Healthy donor | 0.44 | 0 | 0.16 | 0 |  |  |  |
| 95 | Healthy donor | 0.14 | 0 | 0.19 | 0 |  |  |  |
| 96 | Healthy donor | 0.33 | 0 | 0.37 | 0 |  |  |  |
| 97 | Healthy donor | 0.24 | 0 | 0.18 | 0 |  |  |  |
| 98 | Healthy donor | 0.56 | 0 | 0.23 | 0 |  |  |  |
| 99 | Healthy donor | 0.70 | 0 | 0.27 | 0 |  |  |  |
| 100 | Healthy donor | 0.62 | 0 | 0.29 | 0 |  |  |  |
| 101 | Healthy donor | 0.23 | 0 | 0.33 | 0 |  |  |  |
| 102 | Healthy donor | 0.18 | 0 | 0.17 | 0 |  |  |  |
| 103 | Patient under investigation on SARS-CoV-2 PCR negative | 0.71 | 0 | 0.40 | 0 |  |  |  |
| 104 | Patient under investigation on SARS-CoV-2 PCR negative | 0.56 | 0 | 0.13 | 0 |  |  |  |
| 105 | Patient under investigation on SARS-CoV-2 PCR negative | 0.48 | 0 | 0.32 | 0 |  |  |  |
| 106 | Patient under investigation on SARS-CoV-2 PCR negative | 0.38 | 0 | 0.47 | 0 |  |  |  |
| 107 | Patient under investigation on SARS-CoV-2 PCR negative | 0.35 | 0 | 0.28 | 0 |  |  |  |
| 108 | Patient under investigation on SARS-CoV-2 PCR negative | 0.22 | 0 | 0.25 | 0 |  |  |  |
| 109 | Patient under investigation on SARS-CoV-2 PCR negative | 0.21 | 0 | 0.27 | 0 |  |  |  |
| 110 | Patient under investigation on SARS-CoV-2 PCR negative | 0.37 | 0 | 0.27 | 0 |  |  |  |
| 111 | Patient under investigation on SARS- | 0.7 | 0 | 0.25 | 0 |  |  |  |
|  | CoV-2<br>PCR<br>negative |  |  |  |  |  |  |  |
| 11<br>2 | Patient<br>under<br>investigati<br>on SARS-CoV-2<br>PCR<br>negative | 0.61 | 0 | 0.28 | 0 |  |  |  |
| 11<br>3 | Patient<br>under<br>investigati<br>on SARS-CoV-2<br>PCR<br>negative | 0.37 | 0 | 0.37 | 0 |  |  |  |
| 11<br>4 | Patient<br>under<br>investigati<br>on SARS-CoV-2<br>PCR<br>negative | 0.29 | 0 | 0.23 | 0 |  |  |  |
| 11<br>5 | Patient<br>under<br>investigati<br>on SARS-CoV-2<br>PCR<br>negative | 0.47 | 0 | 0.31 | 0 |  |  |  |
| 11<br>6 | Patient<br>under<br>investigati<br>on SARS-CoV-2<br>PCR<br>negative | 1.88 | 1 | 0.56 | 0 |  |  |  |
| 11<br>7 | Patient<br>under<br>investigati<br>on SARS-CoV-2<br>PCR<br>negative | 0.25 | 0 | 0.27 | 0 |  |  |  |
| 11<br>8 | Patient<br>under<br>investigati<br>on SARS-CoV-2<br>PCR<br>negative | 0.23 | 0 | 0.39 | 0 |  |  |  |
| 11<br>9 | Patient<br>under<br>investigati<br>on SARS-CoV-2<br>PCR<br>negative | 0.58 | 0 | 0.32 | 0 |  |  |  |
| 120 | Patient under investigation on SARS-CoV-2 PCR negative | 0.35 | 0 | 0.32 | 0 |  |  |  |
| 121 | Patient under investigation on SARS-CoV-2 PCR negative | 9.49 | 1 | 11.22 | 1 |  |  |  |
| 122 | Patient under investigation on SARS-CoV-2 PCR negative | 6.11 | 1 | 1.37 | 1 | Repeat after 2 weeks | 0.45 | 0.17 |
| 123 | Patient under investigation on SARS-CoV-2 PCR negative | 5.46 | 1 | 0.78 | 0 |  |  |  |
| 124 | Patient under investigation on SARS-CoV-2 PCR negative | 1.31 | 1 | 0.48 | 0 | Repeat after 1 month | 0.25 | 0.18 |
| 125 | Patient under investigation on SARS-CoV-2 PCR negative | 0.91 | 0 | 0.26 | 0 |  |  |  |
| 126 | Patient under investigation on SARS-CoV-2 PCR negative | 0.90 | 0 | 0.41 | 0 |  |  |  |
| 127 | Patient under investigation on SARS-CoV-2 PCR negative | 0.72 | 0 | 0.23 | 0 |  |  |  |
| 128 | Patient under investigation on SARS- | 0.62 | 0 | 0.22 | 0 |  |  |  |
|  | CoV-2<br>PCR<br>negative |  |  |  |  |  |  |  |
| 12<br>9 | Patient<br>under<br>investigati<br>on SARS-CoV-2<br>PCR<br>negative | 0.54 | 0 | 0.21 | 0 |  |  |  |
| 13<br>0 | Patient<br>under<br>investigati<br>on SARS-CoV-2<br>PCR<br>negative | 0.52 | 0 | 0.18 | 0 |  |  |  |
| 13<br>1 | Patient<br>under<br>investigati<br>on SARS-CoV-2<br>PCR<br>negative | 0.50 | 0 | 0.22 | 0 |  |  |  |
| 13<br>2 | Patient<br>under<br>investigati<br>on SARS-CoV-2<br>PCR<br>negative | 0.50 | 0 | 0.20 | 0 |  |  |  |
| 13<br>3 | Patient<br>under<br>investigati<br>on SARS-CoV-2<br>PCR<br>negative | 0.49 | 0 | 0.17 | 0 |  |  |  |
| 13<br>4 | Patient<br>under<br>investigati<br>on SARS-CoV-2<br>PCR<br>negative | 0.47 | 0 | 0.21 | 0 |  |  |  |
| 13<br>5 | Patient<br>under<br>investigati<br>on SARS-CoV-2<br>PCR<br>negative | 0.46 | 0 | 0.41 | 0 |  |  |  |
| 13<br>6 | Patient<br>under<br>investigati<br>on SARS-CoV-2<br>PCR<br>negative | 0.44 | 0 | 0.37 | 0 |  |  |  |
| 13<br>7 | Patient<br>under<br>investigati<br>on SARS-CoV-2<br>PCR<br>negative | 0.44 | 0 | 0.16 | 0 |  |  |  |
| 13<br>8 | Patient<br>under<br>investigati<br>on SARS-CoV-2<br>PCR<br>negative | 0.40 | 0 | 0.26 | 0 |  |  |  |
| 13<br>9 | Patient<br>under<br>investigati<br>on SARS-CoV-2<br>PCR<br>negative | 0.35 | 0 | 0.20 | 0 |  |  |  |
| 14<br>0 | Patient<br>under<br>investigati<br>on SARS-CoV-2<br>PCR<br>negative | 0.33 | 0 | 0.23 | 0 |  |  |  |
| 14<br>1 | Patient<br>under<br>investigati<br>on SARS-CoV-2<br>PCR<br>negative | 0.33 | 0 | 0.20 | 0 |  |  |  |
| 14<br>2 | Patient<br>under<br>investigati<br>on SARS-CoV-2<br>PCR<br>negative | 0.33 | 0 | 0.17 | 0 |  |  |  |
| 14<br>3 | Patient<br>under<br>investigati<br>on SARS-CoV-2<br>PCR<br>negative | 0.30 | 0 | 0.24 | 0 |  |  |  |
| 14<br>4 | Patient<br>under<br>investigati<br>on SARS-CoV-2<br>PCR<br>negative | 0.30 | 0 | 0.28 | 0 |  |  |  |
| 14<br>5 | Patient<br>under<br>investigati<br>on SARS- | 0.29 | 0 | 0.22 | 0 |  |  |  |
|  | CoV-2<br>PCR<br>negative |  |  |  |  |  |  |  |
| 14<br>6 | Patient<br>under<br>investigati<br>on SARS-CoV-2<br>PCR<br>negative | 0.28 | 0 | 0.21 | 0 |  |  |  |
| 14<br>7 | Patient<br>under<br>investigati<br>on SARS-CoV-2<br>PCR<br>negative | 0.22 | 0 | 0.18 | 0 |  |  |  |
| 14<br>8 | Patient<br>under<br>investigati<br>on SARS-CoV-2<br>PCR<br>negative | 0.19 | 0 | 0.22 | 0 |  |  |  |
| 14<br>9 | Patient<br>under<br>investigati<br>on SARS-CoV-2<br>PCR<br>negative | 0.19 | 0 | 0.18 | 0 |  |  |  |
| 15<br>0 | Patient<br>under<br>investigati<br>on SARS-CoV-2<br>PCR<br>negative | 0.18 | 0 | 0.52 | 0 |  |  |  |
| 15<br>1 | Patient<br>under<br>investigati<br>on SARS-CoV-2<br>PCR<br>negative | 0.18 | 0 | 0.18 | 0 |  |  |  |
| 15<br>2 | Dengue<br>IgM/Deng<br>ue IgG | 0.53 | 0 | 0.21 | 0 |  |  |  |
| 15<br>3 | Dengue<br>IgM | 0.42 | 0 | 0.20 | 0 |  |  |  |
| 15<br>4 | Anti-HBS<br>pos | 0.79 | 0 | 0.25 | 0 |  |  |  |
| 15<br>5 | Anti-HBS<br>pos | 0.46 | 0 | 0.20 | 0 |  |  |  |
| 15<br>6 | Treponema<br>l Ab pos | 0.54 | 0 | 0.18 | 0 |  |  |  |
| 15<br>7 | Treponema<br>l Ab pos | 0.62 | 0 | 0.32 | 0 |  |  |  |
| 15<br>8 | Anti-HCV<br>pos | 0.80 | 0 | 0.35 | 0 |  |  |  |
| 159 | Anti-HCV pos | 0.36 | 0 | 0.19 | 0 |  |  |  |
| 160 | HBSAg pos | 0.48 | 0 | 0.16 | 0 |  |  |  |
| 161 | HBSAg pos | 0.30 | 0 | 0.18 | 0 |  |  |  |
| 162 | Mumps IgG pos / Measles IgG pos / VZV IgG pos / HSV IgG pos | 0.67 | 0 | 0.27 | 0 |  |  |  |
| 163 | Measles IgG pos / Rubella IgG pos | 0.47 | 0 | 0.18 | 0 |  |  |  |
| 164 | Rubella IgG pos / EBV IgG pos | 0.79 | 0 | 0.27 | 0 |  |  |  |
| 165 | Mumps G pos | 0.77 | 0 | 0.26 | 0 |  |  |  |
| 166 | VZV IgG pos | 0.61 | 0 | 0.22 | 0 |  |  |  |
| 167 | HSV IgM pos | 0.98 | 0 | 0.25 | 0 |  |  |  |
| 168 | CMV IgG pos / CMV IgM pos | >11 | 1 | 2.11 | 1 |  |  |  |
| 169 | CMV IgG pos / CMV IgM pos | 0.49 | 0 | 0.26 | 0 |  |  |  |
| 170 | EBV IgG pos / EBV IgM pos | 10.84 | 1 | 2.35 | 1 |  |  |  |
| 171 | Rubella IgG pos | 0.52 | 0 | 0.18 | 0 |  |  |  |

**S2 Table:** Summary serological results from 15 studies based on Euroimmun test

| References | Sample Tested | Sensitivity | Control | Specificity | Remarks |
| --- | --- | --- | --- | --- | --- |
| Leaflet package of EUROIMMUN Version: 2020-05-06 | 91 cases (94 samples) | IgA (cut off $\geq 1.1$)<br>$\leq 10$ days (N=66) = 51.5%<br>$>10-20$ days (N=12) = 91.7%<br>$\geq 21$ days (N=16) = 100%<br>IgG (cut off $\geq 1.1$)<br>$\leq 10$ days (N=66) = 30.3%<br>$>10-20$ days (N=12) = 75%<br>$\geq 21$ days (N=16) = 93.8% | 1241 controls (849 blood donors, 90 children, 200 pregnant women, 40 Influenza, 22 EBV & heterophilic Ab, 40 RF) | IgA (cut off $\geq 0.8$) = 88.2%<br>IgA (cut off $\geq 1.1$) = 92.4%<br>IgG (cut off $\geq 0.8$) = 99%<br>IgG (cut off $\geq 1.1$) = 99.6% | - Reference Okba et al., 2020 |
| Okba et al., Emerging Infectious Diseases | Netherlands 3 cases (2 mild, 1 severe), 10 serum samples | IgA = 70% (7/10)<br>IgG = 60% (6/10) | 203 controls (45 blood donors, 5 Adenovirus, 2 Bocavirus, 2 Enterovirus, 9 HMPV, 13 Influenza A, 6 Influenza B, 9 Rhinovirus, 9 RSV, 4 PIV-1, 4 PIV-3, 1 Mycoplasma pneumoniae, 5 CMV, 7 EBV, 19 229E, 18 NL63, 38 OC43, 7 MERS-CoV, 2 SARS-CoV) | IgA (cut off $\geq 0.9$) = 94.6% (192/203)<br>IgG (cut off $\geq 0.3$) = 96% (195/203) | - Beta-version of EUROIMMUN using in-house cutoff value based on the mean background<br>- Severe case developed Ab sooner and in higher concentrations.<br>- Cross-reactivity with HCoV-OC43, SARS-CoV<br>- Correlation with plaque reduction neutralization assay (PRNT<sub>50</sub>) r =0.5-0.9 |
| | Berlin 9 cases, 31 serum samples | IgA = 88.9% (8/9)<br>IgG = 88.9% (8/9) | 24 serum samples (4 HCoV-229E, 3 HCoV-HKU1, 7 HCoV-NL634 HCoV-OC43, 3 MERS-CoV, 3 SARS-CoV) | IgA (cut off $\geq 0.9$) = 87.5% (21/24)<br>IgG (cut off $\geq 0.3$) = 87.5% (21/24) | |
| Jassskelainen et al., Euro Surveill. | Finland 39 cases (9 mild, 15 moderate, 13 severe, 3 unknown) | Median time after onset of symptoms<br>IgA = 11 days (5-20 days)<br>IgG = 12 days (5-20 days) | 37 Cross-reactivity panel: Influenza, Parainfluenza, RSV, Enterovirus, Coronavirus OC43, NL63, 229E | IgA (cut off $\geq 1.1$) = 73%<br>IgG (cut off $\geq 1.1$) = 91.9% | - No correlation with severity was clearly seen.<br>- IgA cross react with HCoV-OC43 and other respiratory viruses e.g., |
|  |  |  |  |  | Influenza, Parainfluenza, RSV, Adenovirus<br>- IgA was not suggested for initial screening due to low specificity<br>- IgG cross react with HCoV-OC43<br>- one case with mild disease severity still has IgG negative 16 days after onset of symptoms |
| Jassskelainen et al., Journal of Clinical Virology | Finland 62 cases (70 samples) | IgA (cut off $\geq 1.1$) 87.8%<br>IgG (cut off $\geq 1.1$) 70.7% | 81 controls (Autoimmune and respiratory virus in 2018 and 2019) | IgA (cut off $\geq 1.1$) 68.3%<br>IgG (cut off $\geq 1.1$) 86.8% | - No correlation between disease severity (N=55) and microneutralization titers<br>- IgA has 87.8% sensitivity and IgG has 70.7% sensitivity compared with Microneutralization Test<br>- No cross-react with RF |
| Montesinos et al., Journal of Clinical Virology | Belgium 128 cases | IgA (cut off $\geq 0.8$)<br>Overall = 83.6%<br><7 days (N=29) = 65.5%<br>8-14 days (N=62) = 87.09%<br>>15 days (N=33) = 93.93%<br>IgG (cut off $\geq 0.8$)<br>Overall = 61.7%<br><7 days (N=29) = 17.2%<br>8-14 days (N=62) = 66.12%<br>>15 days (N=33) = 90.9% | 72 controls (5 EBV, 11 CMV, 8 M.pneumoniae, 1 Parvovirus, 1 HBV, 1 Bartonella henselae, 1 Brucella spp, 3 autoimmune) | IgA (cut off $\geq 0.8$) = 86.1%<br>IgG (cut off $\geq 0.8$) = 98.6% | - IgA cross-react with EBV, M. pneumoniae, Anti-PL12 and also with healthy control without any known confounding factors. |
| Tang et al., American Association for Clinical Chemistry | USA 48 cases (103 samples) Hospitalized with multiple comorbidities | IgG (cut off $\geq 0.8$)<br>< 3 days (N=12) = 0%<br>3-7 days (N=20) = 25%<br>8-13 days (N=23) = 56.5%<br>$\geq 14$ days (N=48) = 85.4% | 153 controls (50 serum in 2015, 80 symptomatic but PCR negative for SARS-CoV-2, 5 other coronaviruses, HKU1, NL63, 229E, 4 influenza, 5 CMV, 3 EBV VCA IgG, 3 EBV VCA IgM, 2 EBV IgM and IgG, 1 RF) | IgG (cut off $\geq 1.1$) = 96.7%<br>IgG (cut off $\geq 0.8$) = 94.8% | - 3 of the false positives were from sample collected in 2015.<br>- 3 patients had no Ab responses at > 14 days post-infection |
| Elslande et al.,<br>Clinical Microbiology and Infection | Belgium 94 cases (167 samples) | IgG (cut off $\geq 1.1$)<br>14-25 days = 89.5% | 103 controls (49 respiratory infection, 14 PCR negative for SARS-CoV-2, 40 other pathogens e.g., CMV, EBV, HIV) | IgA (cut off $\geq 1.1$) = 73.8%<br>IgG (cut off $\geq 1.1$) = 96.1% | - IgA cross-react with a number of other infections<br>- Due to low specificity of IgA, not recommend using IgA for screening of asymptomatic persons<br>- IgG cross react with EBV, CMV, Enterovirus/Rhinovirus+, HSV1+, Streptococcus pneumoniae+ |
| Kruttgen et al.,<br>Journal of Clinical Virology | Germany 31 cases (50 samples) | IgG (cut off $\geq 1.1$)<br>7-17 days = 86.4% | 25 controls (PCR negative for SARS-CoV-2) | IgG (cut off $\geq 1.1$) = 96.2% | |
| Hardy et al.,<br>Clin Chem Lab Med | Belgium 44 cases | IgG (cut off $\geq 1.1$) = 96% | 81 controls (1 NL63, 1 OC43, 7 HBV, 3 HAV, 1 Adenovirus, 1 HSV and CMV, 8 CMV, 5 Parvovirus B19, 1 HIV, 4 antistreptolysin O (ASLO), 1 anti Treponema pallidum, 1 Borrelia, 10 Mycoplasma pneumoniae, 16 Toxoplasma gondii, 1 RF, 7 anti-TPO, 4 agglutinins, 1 direct coombs, 1 high level of IgM, 1 high IgA, 6 healthy) | IgG (cut off $\geq 1.1$) = 98% | - Cross-react with anti-TPO, anti-HAV, ASLO |
| Beavis et al.,<br>Journal of Clinical Virology | USA 82 cases | IgA (cut off $\geq 1.1$) = 82.9%<br>$\geq 4$ days after positive PCR 90.5%<br>IgG (cut off $\geq 1.1$) = 67.1%<br>$\geq 4$ days after positive PCR 100% | 86 controls | IgA (cut off $\geq 1.1$) = 88.4%<br>IgG (cut off $\geq 1.1$) = 97.7% | - Borderline cross-react with NL63 and OC43 |
| Nicol et al.,<br>Journal of Clinical Virology | France 82 cases (141 samples) | IgA (cut off $\geq 1.1$)<br>Overall = 86.7%<br>$\leq 7$ days (N=56) = 59.4<br>8-14 days (N=44) = 79.3% | 155 controls (50 SARS-CoV-2 RT-PCR negative, 50 control before March 2019, | IgA (cut off $\geq 1.1$) = 82.7%<br>IgG (cut off $\geq 1.1$) = 96.7% | - IgA and IgG cross-react with pregnant serum, RF, patients with symptoms of pneumonia |
| | | $\geq 15$ days (N=98) = 100%<br>IgG (cut off $\geq 0.8$)<br>Overall = 78.3%<br>$\leq 7$ days (N=56) = 28.1%<br>8-14 days (N=44) = 72.4%<br>$\geq 15$ days (N=98) = 100% | 25 cross-reactivity panel, 10 pregnant women, 10 RF) | | |
| Kohmer et al.,<br>Journal of Medical Virology | Germany 33 cases | IgG (cut off $\geq 1.1$)<br>5-9 days (N=17) = 58.8%<br>10-18 days (N=16) = 93.8% | 21 controls (SARS-CoV, OC43, HKU1, NL63, 229E, EBV, CMV) | IgG (cut off $\geq 1.1$) = 96.2% | - All IgG positive tested samples showed neutralizing properties in the PRNT (tier $\geq 1.20$) |
| Kohmer et al.,<br>Journal of Clinical Virology | Germany 45 cases | IgG (cut off $\geq 1.1$) = 71.1% | 22 controls (SARS-CoV, OC43, HKU1, NL63, 229E, EBV, CMV) | IgG (cut off $\geq 1.1$) = 100% | - Equivocal results with OC43 and negative control cohort)<br>- Correlate well with PRNT |
| Theel et al.,<br>Journal of Clinical Microbiology | USA 56 cases (224 samples from 33 in-patient and 23 out-patient) | IgG (cut off $\geq 1.1$)<br>$< 7$ day (N=38) = 0 (in-patient), (N=11) = 18.2% (out-patient)<br>8-14 days (N=91) = 27.5% (in-patient)<br>$> 15$ days (N=61) = 100% (in-patient)<br>$> 20$ days (N=23) = 91.3% (out-patient) | 149 controls from 2018<br>105 cross-reactivity panel (CMV, Influenza, Mycoplasma pneumoniae, Chlamydia pneumoniae, Streptococcus pneumoniae urinary antigen, coronavirus, metapneumovirus, RSV, adenovirus, rhinovirus/enterovirus, HBV, HCV, HIV) | IgG (cut off $\geq 1.1$)<br>99.3% (healthy)<br>96.2% (Cross-reactivity panel)<br>98% (overall) | |
| Tuaillon et al.,<br>Journal of Infection | France 38 cases | IgA (cut off $\geq 1.1$) 93.3%<br>IgG (cut off $\geq 1.1$) 93.3% | 20 controls | IgA (cut off $\geq 1.1$) 80%<br>IgG (cut off $\geq 1.1$) 85% | |

